# Single genome amplification and molecular cloning of HIV-1 populations in acute HIV-1 infection: implications for studies on HIV-1 diversity and evolutionary rate

**DOI:** 10.1101/2025.03.17.25324117

**Authors:** Anthony Y.Y. Hsieh, Amin S. Hassan, Jamirah Nazziwa, Sara Karlson, Lovisa Lindquist, Jonathan Hare, Anatoli Kamali, Etienne Karita, William Kilembe, Matt A. Price, Per Björkman, Pontiano Kaleebu, Susan Allen, Eric Hunter, Jill Gilmour, Sarah Rowland-Jones, Eduard J. Sanders, Joakim Esbjörnsson

## Abstract

**Background:** Human immunodeficiency virus type 1 (HIV-1) is one of the fastest evolving human pathogens. Understanding transmission, within-host adaptation, and evolutionary dynamics are pivotal for development of interventions and vaccines. HIV-1 infection is generally caused by one single transmitted founder virus (TFV), and TFV sequences have typically been obtained using single genome amplification (SGA). However, suboptimal sample quality can result in sequencing failures, representing non-trivial losses considering the scarcity of acute HIV-1 infection (AHI) samples. Sequencing failures may be mitigated by molecular cloning (MC), a method that can be less vulnerable to sample quality but more susceptible to PCR errors. Here, we explore the feasibility of supplementing SGA with MC data using samples from clinical and research cohorts to determine whether sequence diversity and evolutionary rate estimates are comparable between the two techniques.

**Methods:** Participants were enrolled in an East African research cohort from the International AIDS Vaccine Initiative 2006-2011 or a clinical cohort from Sweden (1983-2011). SGA and MC sequencing were done on the HIV-1 *env V1-V3* region (approximately 940 base pairs). Within-host sequence diversity was determined from maximum likelihood phylogenetic trees and evolutionary rate by Bayesian phylogenetic analysis. Highlighter and Poisson-Fitter tools, Hamming distances, and assessment of star phylogenies were used to quantify TFVs.

**Results:** Participants with AHI (N=100, median age 30.3 years, 15% female) were included, contributing 350 samples from four longitudinal time points 10-540 days post infection. SGA succeeded on 90% of research cohort and 48% of clinical cohort samples. Comparative analysis of linked SGA and MC data from 10 samples indicated that approximately eight sequences were necessary for diversity estimates. Consistently higher sequence diversity was observed among MC relative to SGA sequences (mean±SD 0.009±0.007 and 0.006±0.006 substitutions/site, p<0.001), whereas evolutionary rates were similar between the two methods (mean±SD 0.014±0.006 vs. 0.014±0.009 substitutions/site/year, p=0.673). Five participants with visits within 45 days post infection were eligible for TFV quantification and all found to have one TFV using both MC and SGA data.

**Conclusion:** MC data is a suitable supplement for SGA-based studies to preserve the value of precious samples for evolutionary rate but not sequence diversity analysis.

## INTRODUCTION

Despite a heterogenous human immunodeficiency virus type 1 (HIV-1) population seen in chronic infection, newly infected recipients mostly present with a genetically homogenous virus population during acute HIV-1 infection (AHI)^1,2^. This “bottleneck” effect during HIV-1 transmission is typically a result of a single transmitted founder virus (TFV) infection^3–8^. Selection of the TFV is an elaborate confluence of route of infection, viral fitness, and host immunity^8–10^ and over time HIV-1 genetic diversity within the host accumulates due to genetic drift and selection until it stabilizes during chronic infection^11–14^. Over the past two decades, characterizing the evolution of HIV-1 into a highly heterogenous population during chronic infection and studying sequence diversity in people with chronic HIV-1 have become important fields of study with implications for virus control, vaccine design, and cure research^4,5,7,15–18^. Taken together, the continual study of intra-host virus diversity in different populations and settings remains a priority, particularly in AHI, where it remains to be fully characterised.

Early studies to characterize intra-host virus diversity used molecular cloning (MC) of PCR-amplified HIV-1 genes to isolate and determine individual HIV-1 sequences^2,3,19,20^. However, this method is susceptible to *Taq* polymerase errors, such as template switching, which may lead to cloning and sequencing of recombinant amplicons^21,22^, thus overestimating diversity estimates and biasing evolutionary analysis. It is also possible that original template sequences are not proportionately amplified prior to MC, and thereby non-reflective of quasi-species composition^23^ and resulting in errors in diversity estimates.

Despite the popularity of next generation sequencing approaches, the current gold standard for measuring intra-host virus diversity is single-genome amplification (SGA), which involves isolating individual HIV-1 genomes through limiting dilution^6,24^. Importantly, selection of individual sequences by limiting dilution occurs before PCR amplification, and thus any potential PCR error would manifest within reads of a sequenced template. In contrast, PCR errors in MC accumulate prior to the selection of clones and would be dispersed across multiple sequenced templates. Therefore, SGA PCR errors can be identified and filtered out in the analysis pipeline, whereas MC PCR errors may be misattributed as diversity in the original sample.

Despite the superior accuracy of the SGA method, by amplifying the starting material prior to selection of clones, the MC method may be more likely to generate enough sequences for diversity and evolutionary rate analysis in circumstances where starting material is low and/or sample quality is suboptimal. Given that intra-host diversity studies are often characterized from comparatively scarce acute HIV-1 samples, instances in which SGA fails to generate sufficient sequences for analysis represent a non-trivial loss. Thus, for studies in which achieving adequate statistical power is hampered by the rarity of samples, it may be worthwhile to recover these sequences using MC. This is especially true given that a costly approach of many more single genomes would have to be amplified to match the sensitivity of the MC method.

In this study, we explored the feasibility of supplementing SGA with MC data as part of a large research cohort from the International AIDS Vaccine Initiative (IAVI) and a historical clinical cohort from Sweden^25–27^. We compared the estimated number of TFVs, HIV-1 sequence diversity, and HIV-1 evolutionary rate, as measured from sequences generated by these two techniques, respectively. We also assess the number of sequences necessary for diversity measurements for both methods. We hypothesized that sequence diversity would be higher when measured using MC compared to SGA due to errors inherent to the former methodology, but that the within-host evolutionary rate would be comparable between sequences generated by the two techniques. We also assessed the utility of qPCR quantification of HIV-1 genomes as an additional step in the SGA workflow, both to inform the limiting dilution and to use its concordance with clinically determined HIV-1 plasma viral load (pVL) as an indicator of sample quality.

## METHODS

### Study participants

Study participants were selected based on our work described elsewhere^25–27^. Briefly, participants were adults (≥18.0 years old) enrolled either in a research cohort (IAVI protocol C)^28^ 2006-2011 from sites in Kilifi, Kenya; Kigali, Rwanda; Masaka, Uganda; and Lusaka, Zambia; or in a routine historical clinical cohort at the Skåne University Hospital in Lund, Sweden prior 1983-2011. Eligibility included participants with AHI, as defined by samples collected at either Fiebig stage I (HIV-1 RNA positive) or Fiebig stage II (HIV-1 p24 antigen positive but with a negative HIV-1 antibody test)^29^. Plasma samples from four longitudinally matched time points were obtained based on the number of days from the estimated date of infection (EDI) as follows: Time point I (10-14 days), time point II (30±15 days), time point III (90±30 days), and time point IV (360±180 days). Plasma samples from the research and clinical cohorts were stored at −80°C and −20°C, respectively. Participants enrolled in the research cohort provided written informed consent and ethics approvals were obtained from ethics review boards of each participating country^28^. Approvals were obtained from ethics boards at each site, including the Kenyatta National Hospital Ethical Review Committee of the University of Nairobi, Rwanda National Ethics Committee, Uganda Virus Research Institute Science and Ethics Committee, Uganda National Council of Science and Technology, University of Zambia Research Ethics Committee, and Emory University Institutional Review Board^28,30^. Ethics approval for the clinical cohort was obtained from the Lund University Ethical Review Board, Sweden (Dnr 2013/772).

### Single genome amplification and sequencing

SGA and sequencing was done as previously described^13,26,31,32^. Briefly, archived plasma samples were retrieved, thawed, and 100 µl aliquots used for HIV-1 RNA extraction using the RNeasy lipid tissue Mini Kit as per manufacturer’s instructions with minor modifications^32^ (Cat# 74804, Qiagen). Electron microscopy-quantified HIV-1 virions (Cat# 10-118-000, Advanced Biotechnologies Inc) spiked in PBS and neat PBS were used as positive and negative controls, respectively. A two-step RT-PCR protocol was used to amplify HIV-1 genomes. In the first step, the samples were reverse transcribed using random hexamers (Cat# N8080127, Thermo Fisher Scientific) to generate cDNA templates using the SuperScript^TM^ IV Reverse Transcriptase Kit according to manufacturer’s instructions (Cat# 18090010, Thermo Fisher Scientific). Unlike conventional SGA workflows, a qPCR step was introduced. Specifically, the cDNA templates were quantified by qPCR using the SYBR^TM^ Select Master Mix kit as per manufacturer’s instructions (Cat# 44-729-08, Applied Biosystems). The qPCR results were used to inform calculations for serial limited dilutions of cDNA templates, aiming at 0.4 copies/µl (one copy template in 2.5 µl) input for SGA. In the second step, diluted cDNA templates were used for outer and nested PCR using gene-specific primers targeting the HIV-1 *env V1-V3* region (approximately 940 base pairs, nucleotides 6430-7374 in HXB2; GenBank accession number K03455). Primers JE12F (forward) and V3A_R2 (reverse), primers E20A_F (forward) and JA169_R (reverse) were used for outer and nested PCR, respectively^33^. All PCR reactions were done using the DreamTaq Green DNA Polymerase kit as per manufacturer’s instructions (Cat# EP0712, Thermo Fisher Scientific). Nested PCR products were visualized using agarose gel electrophoresis, and successful amplificons were retrieved, purified and sequenced by the BigDye Terminator Cycle Sequencing Kit, using the primers E20A_F and JA169_R according to manufacturer’s instructions (Applied Biosystems). Twenty-four SGAs were targeted for sequencing for each sample.

### Molecular cloning and sequencing

Molecular cloning (MC) and sequencing were done as previously described^32^. Briefly, the HIV-1 *V1-V3 env* region (as above) was amplified from 5 μl of the extracted HIV-1 RNA eluate that was used for SGA. Specifically, the outer primers used in the SGA approach (JE12F and V3A_R2) was used for one-step RT-PCR (SuperScript™ III One-Step RT-PCR System with Platinum® Taq DNA Polymerase, ThermoFisher Scientific); and the nested primers used above (E20A_F and JA169) were used for nested PCR (DreamTaq DNA Polymerase, ThermoFisher Scientific), as previously described^32,34^. The amplified *V1-V3* region was then cloned using the TOPO™ TA Cloning™ Kit with One Shot™ TOP10 Chemically Competent *E. coli* (ThermoFisher Scientific) according to the manufacturer’s instructions. Twenty-three colonies were routinely picked from each sample and amplified with DreamTaq DNA Polymerase (ThermoFisher Scientific) using conventional M13 primers (−20 and −24). Individual clones were purified (MinElute PCR Purification Kit, Qiagen), and sequenced by the BigDye Terminator Cycle Sequencing Kit (Applied Biosystems), using the primers E20A_F and JA169_R, according to the manufacturers’ instructions.

### Sequence data management

An automated workflow was set up in Geneious Prime (https://www.geneious.com) for sequence data management. Briefly, forward and reverse sequence reads were compiled, poor-quality ends trimmed, and *de novo* assembly done using default settings. Assembled contigs were mapped to the HXB2 reference sequence (GenBank accession number K03455), prior to the generation of a global alignment using the Clustal algorithm. A Neighbour Joining (NJ) phylogenetic tree was explored for potential sample mix-up, sample mislabelling and contamination. Sequences suggestive of a sample mix-up, mislabelling or contamination were excluded from further analysis. The pairwise homoplasy index (PHI) test was applied together with an in-house Perl script to iteratively and exhaustively screen for putative recombinants, as previously described^35,36^. Sequences suggestive as putative recombinants were excluded from further analysis. The remaining sequence alignment was used in downstream analyses.

### Analysis of transmitted founder viruses

Sequences generated from time point I or time point II were used for the determination of transmitted founder viruses (TFVs), as described^4^. Briefly, SGA or MC sequences were aligned per time point in Geneious Prime using the Clustal algorithm with default settings. The number of TFVs were then determined using a three-pronged approach. First, the sequence alignments were visually inspected using the Highlighter tool (HIV Sequence Database, National Institutes of Health, https://www.hiv.lanl.gov/content/sequence/HIGHLIGHT/highlighter_top.html). Homogeneously distributed alignments were considered single TFV infections. Depending on the heterogeneity of the distribution, alignments sharing two, three or more nucleotide substitution sites were considered as two, three or more TFV infections, respectively. Second, the sequence alignments were used to estimate pairwise hamming distances (HDs), to explore its frequency distribution, mean of best fitting distribution, and goodness of fit p-values using the Poisson-Fitter tool (HIV Sequence Database, National Institutes of Health, https://www.hiv.lanl.gov/content/sequence/POISSON_FITTER/poisson_fitter.html). Graphical distribution of the HDs with one, two, three or more peaks and with supporting statistics were considered single, two, three or more TFV infections, respectively. Third, the sequence alignments were used to generate NJ phylogenies, which were further explored in unrooted radial layout. Star-shaped phylogenies (with a single node suggesting a monophyletic lineage) were considered single TFV infections. Bifurcated phylogenies (with two, three or more nodes, suggesting a polyphyletic lineage) were considered two, three or more TFV infections, respectively.

### Analysis of within-host diversity

The sequence alignments from each time point were used to generate 200 bootstrapped maximum likelihood (ML) phylogenies using the Genetic Algorithm for Rapid Likelihood Inference (GARLI, Evolution and Genomics, https://evomics.org/resources/software/molecular-evolution-software/garli/). An in-house Perl script was then used to extract the within-host genetic diversity for each time point by averaging pairwise tree distances between sequences obtained from the same sample time point, as previously described^13^. To assesses the number of sequences needed for HIV-1 diversity estimation, the influence of the sequence abundance on the estimated diversity was explored. Participants with at least 15 SGA and MC sequences, respectively, were eligible. Two sequences were randomly selected from each sample and used to construct an ML tree in IQ-TREE from which the diversity estimate was extracted using the same Perl script as above. This was then iterated 100 times for each sample, resulting in 100 sample-specific diversity estimates based on two randomly selected samples. Next, these steps were independently repeated by adding one randomly selected sequence for each step until 15 sequences had been randomly selected, and 15 × 100 diversity estimates generated for each sample.

### Analysis of within-host evolution

Participant-level SGA or MC sequences across all time points were aligned as described above and used to generate within-host evolutionary rate estimates. The Bayesian Evolution Analysis Utility (BEAUti) was used to set up .xml files for sequence analysis in Bayesian Evolutionary Analysis by Sampling Trees (BEAST). Nucleotide substitution rates were estimated using the HKY substitution model with estimated base frequencies, four gamma categories and two data partitions into codon positions. Furthermore, a strict clock model with a constant coalescent population size was set as tree prior. Each analysis was run for a 100 million Markov Chain Monte Carlo (MCMC) iterations, with sampling done after every 10,000 iterations. Log files were analysed in Tracer, with Effective Sample Sizes (ESSs) >100 reflecting sufficient posterior distributions of model parameters.

### Statistical analysis

Participant demographic data were presented with summary statistics. Continuous data were presented using medians and interquartile ranges (IQRs), while categorical data were presented using frequencies and percentages. Based on data distributions, chi-squared and Mann-Whitney U tests were done to compare sample quality between the research and clinical cohorts, and Pearson’s correlations were used to characterize the concordance between the quantity of plasma HIV-1 viral load (HIV-1 pVL) as historically determined from the research site/clinic and during the current sequencing from the HIV-1 qPCR step introduced in this study. Cochran–Armitage test for trend was used to compare recombinant sequences with time point. Within-host diversity and evolutionary rates were compared between SGA and MC methods using paired and unpaired t-tests within and between participants, respectively.

## RESULTS

### Characteristics of study participants

Overall, 100 participants were eligible for SGA sequencing (median age, 30.3 [IQR, 24.3-36.1] years and male [n=85, 85%], Table 1). Of these, 74 participants were from the research cohort and 26 participants from the clinical cohort. Overall, 50 samples were missing from 39 participants, resulting in 350 available samples in total (Figure S1). All participants had at least one sample available.

**Table 1.** Characteristics of participants with acute HIV-1 infection from the clinical and research cohorts in this study.

| Baseline variables |  | Female (n=15) | Male (n=85) | Overall (n=100) |
| --- | --- | --- | --- | --- |
| Age (years) | Median (IQR) | 28.9 (25.6 – 33.5) | 30.7 (24.2 – 36.6) | 30.3 (24.3 – 36.1) |
| Age group (years) | 18.0 – 24.9 | 3 (20.0) | 24 (28.2) | 27 (27.0) |
|  | 25.0 – 34.9 | 10 (66.7) | 36 (42.4) | 46 (46.0) |
|  | 35.0+ | 2 (13.3) | 25 (29.4) | 27 (27.0) |
| Year of infection | <2009 | 6 (40.0) | 36 (42.4) | 42 (42.0) |
|  | 2009 – 2010 | 8 (53.3) | 33 (38.8) | 41 (41.0) |
|  | 2011+ | 1 (6.7) | 16 (18.8) | 17 (17.0) |
| Risk group | HET | 15 (100.0) | 36 (42.4) | 51 (51.0) |
|  | MSM | 0 (0.0) | 49 (57.6) | 49 (49.0) |
| Country | Rwanda | 6 (40.0) | 8 (9.4) | 14 (14.0) |
|  | Uganda | 4 (26.7) | 9 (10.6) | 13 (13.0) |
|  | Kenya | 3 (20.0) | 29 (34.1) | 32 (32.0) |
|  | Zambia | 2 (13.3) | 13 (15.3) | 15 (15.0) |
|  | Sweden | 0 (0.0) | 26 (30.6) | 26 (26.0) |
| HIV-1 subtype | A1 | 10 (66.7) | 37 (43.5) | 47 (47.0) |
|  | B | 0 (0.0) | 8 (9.4) | 8 (8.0) |
|  | C | 2 (13.3) | 17 (20.0) | 19 (19.0) |
|  | D | 3 (20.0) | 3 (3.5) | 6 (6.0) |
|  | Others* | 0 (0.0) | 6 (7.1) | 6 (6.0) |
|  | Missing | 0 (0.0) | 14 (16.5) | 14 (14.0) |
| Transmitted founder viruses | Single | 10 (66.7) | 54 (63.5) | 64 (64.0) |
|  | Multiple | 3 (20.0) | 16 (18.8) | 19 (19.0) |
|  | Missing | 2 (13.3) | 15 (17.7) | 17 (17.0) |

\*Others: HIV-1 subtypes F1 (n=1), G (n=1) and recombinants A2D (n=1), AE (n=2), BG (n=1).
4 Clinical cohort recruited from Kenya, Rwanda, Uganda, and Zambia, and research cohort recruited from Sweden. Abbreviations: IQR (interquartile
5 ranges), HET (heterosexual) and MSM (men who have sex with men).

### qPCR measurements for limiting dilution and sample quality estimates

SGA relies on limiting dilution to isolate HIV-1 single genomes, and the dilution itself was informed by quantifying the cDNA using qPCR. The qPCR data was also used to evaluate sample quality by comparing with the visit-specific HIV-1 pVL. HIV-1 pVL and qPCR data were available for 282 longitudinal visits from 92 participants. Of these, the correlation between HIV-1 pVL and qPCR was R^2^=0.44 (p<0.001) for research samples and R^2^=0.29 (p<0.001) for clinical samples (Figure S2A-B). Among the 277 visits with detectable HIV-1 pVL, 57 (21%) could not be detected by qPCR (Figure S3). This was higher in the clinical cohort than in the research cohort (27 [66%] vs. 30 [13%] samples, p<0.001), and was associated with older sample collection date (median difference 535 days, p=0.008). In sensitivity analyses excluding samples collected before 2005, the effect of sample age could not be detected in either clinical (p=0.14) or research (p=0.15) cohorts. When the analysis was restricted to those with detectable HIV-1 qPCR, the correlation with historic HIV-1 pVL improved to R^2^=0.62 (p<0.001) and R^2^=0.48 (p=0.006) for the research and clinical participants, respectively (Figure S2C-D).

### Single genome amplification and molecular cloning

From the 350 available samples, 50 samples from 29 participants could not be PCR amplified and therefore not sequenced. Of the remaining 300 samples successfully amplified and sequenced, 14 samples from 13 participants yielded no sequence data (Figure S1). Among available samples, PCR amplification or sequencing failures were 52% in the clinical cohort and 10% in the research cohort. In total, 286 samples from 92 participants yielded sequencing data, however, 18 (6%) samples from 15 participants were excluded from further analysis because of potential contamination (Figure 1A). Finally, sequence data from 268 (94%) samples from 86 participants were included in downstream analyses (Figure S1). Each sample had a mean±standard deviation (SD) of 16.1±5.9 sequences. The number of sequences obtained per sample was higher in the research cohort compared to the clinical cohort (16.5±5.7 sequences vs. 13.5±6.7 sequences, p=0.005). There was no relationship between sample collection date and number of sequences obtained per sample (p=0.17).

**Figure 1.**
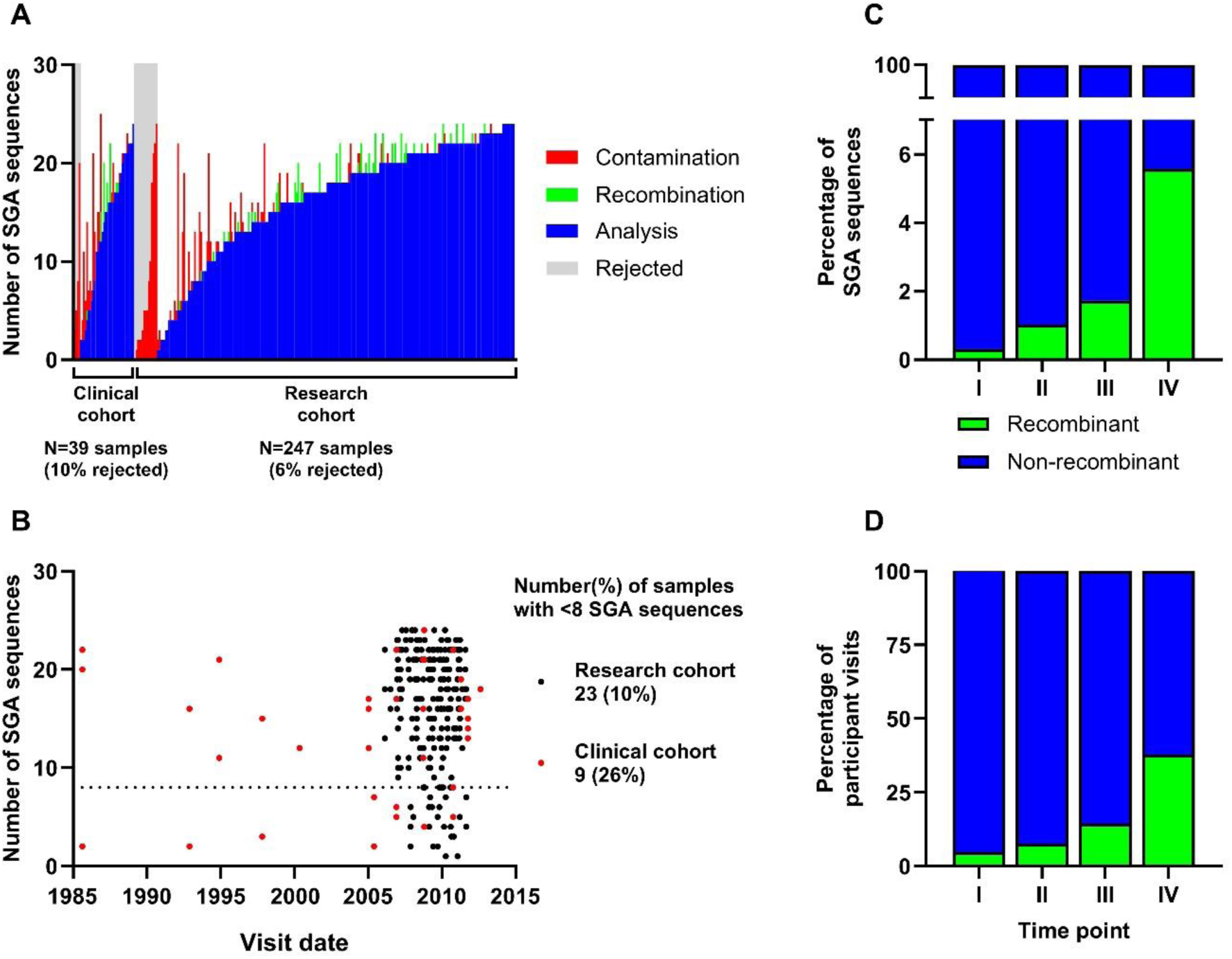
Summary of SGA sequences. (A) Proportions of SGA sequences that were analyzed (blue), identified as recombinant (green), or rejected (gray) due to contamination (red) are shown. (B) Relationship between visit date and number of sequences obtained per sample in research (black dots) and clinical (red dots) samples. (C) Frequency of recombinant sequences among all obtained sequences and (D) frequency of samples with recombinant sequences at each time point. Abbreviations: SGA (single genome amplification).

There was a total of 4391 SGA sequences. Of these, putative PCR-induced recombination was detected in 94 (2.1%) sequences. The proportion of putative recombinant sequences per total number of sequences increased with later time point, ranging from 0.3% in time point I to 5.6% in time point IV (p=0.011, Figure 1C). When grouped by sample, the prevalence of samples containing a putative recombinant sequence followed a similar pattern, in which 4.7% of time point I samples had recombinant sequences and 37.8% of time point IV participants had recombinant sequences (p<0.001, Figure 1D).

### The number of sequences needed for HIV-1 diversity estimation

The median diversity ranged from 0.001 [IQR, 0.000-0.003] to 0.003 [0.002-0.003] substitutions/site for samples generated using SGA, and from 0.004 [0.002-0.006] to 0.008 [0.007-0.008] substitutions/site for samples generated using MC. The median diversity increased with the number of sequences until reaching a plateau at approximately eight sequences (Figure 2A).

**Figure 2.**
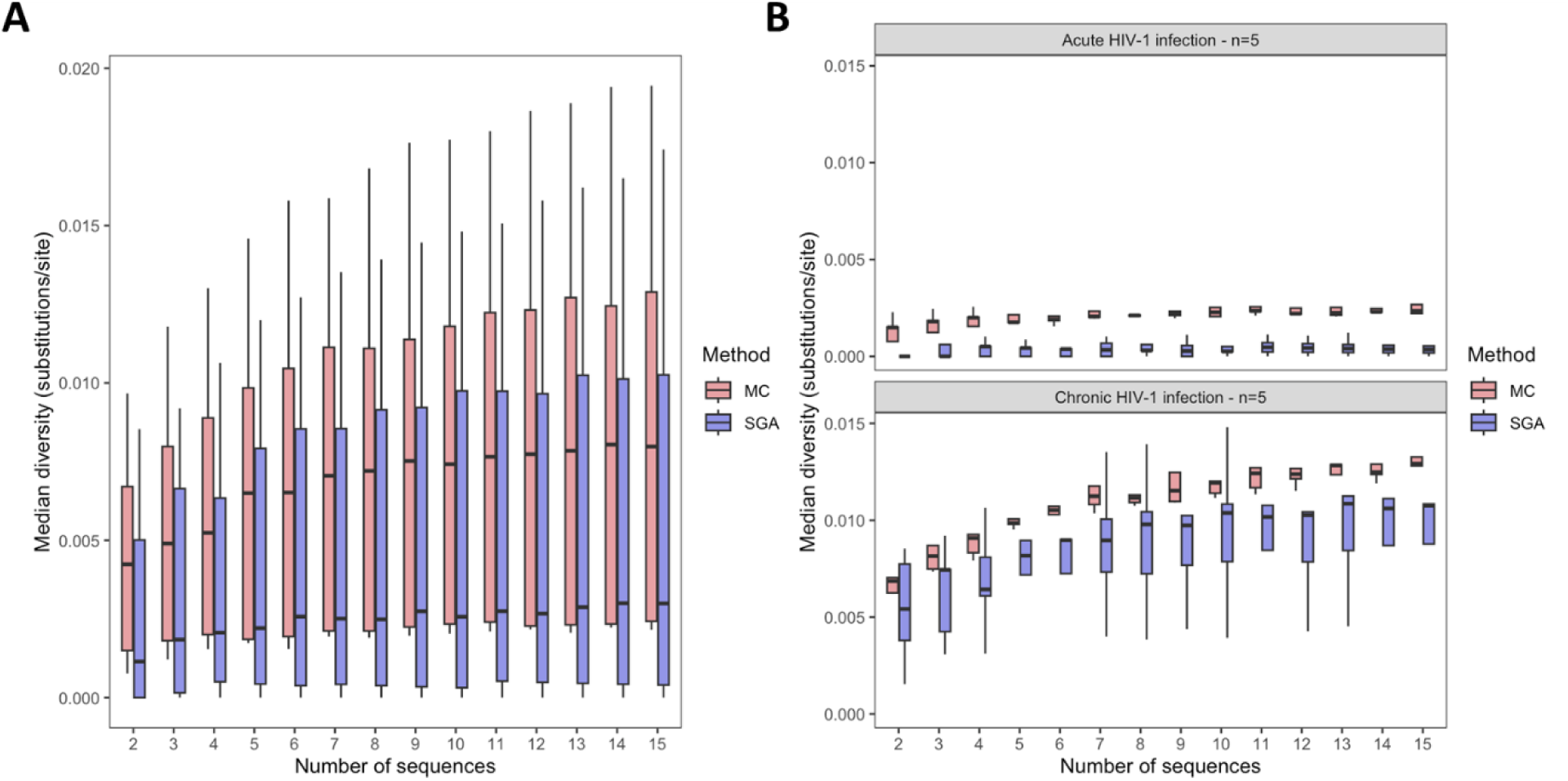
Number of sequences necessary for diversity estimate. (A) Median diversity calculated using 2 to 15 sequences from 10 participants with at least 15 sequences. (B) Participants have been separated between acute (time point I or II) and chronic (time point III or IV), showing diversity estimates depending on number of sequences used. Molecular cloning (red) and single genome amplification (blue). Abbreviations: MC (molecular cloning), SGA (single genome amplification).

When limiting the analysis to the acute HIV-1 infection (time points I and II), diversity estimates were relatively low, with little influence of the number of sequences used, regardless of sequencing approach (Figure 2B). During the chronic infection (time points III and IV), the diversity increased with the number of sequences and plateaued at approximately eight sequences (Figure 2B). The number of sequences to plateau did not seem to differ based on the sequencing approach, however, the diversity estimates obtained from MC sequences were consistently higher than the estimates based on SGA sequences. Among SGA data, and 23 (10%) of research samples and 9 (26%) of routine clinic samples did not reach the threshold of eight sequences and were not considered for comparison of diversity of SGA and MC (Figure 1B).

### Comparison of diversity between SGA and MC

To compare measurements of diversity between SGA and MC methods, 10 participants were selected semi-randomly by ensuring an even distribution across study sites, time points and SGA sequences (Table 2). SGA yielded a mean±SD of 17.7±2.4 sequences for analysis, compared to a 20.2±2.4 sequences from MC. Moreover, *prima facie* of the NJ phylogeny indicated that MC and SGA sequences from the same participant and time point clustered together, time point I sequences had more homogeneous sequences with shorter branch lengths compared to those from later time points, and MC sequences generally had longer branch lengths compared to SGA sequences regardless of time point, suggesting higher diversity in MC compared to SGA sequences (Figure 3).

**Figure 3.**
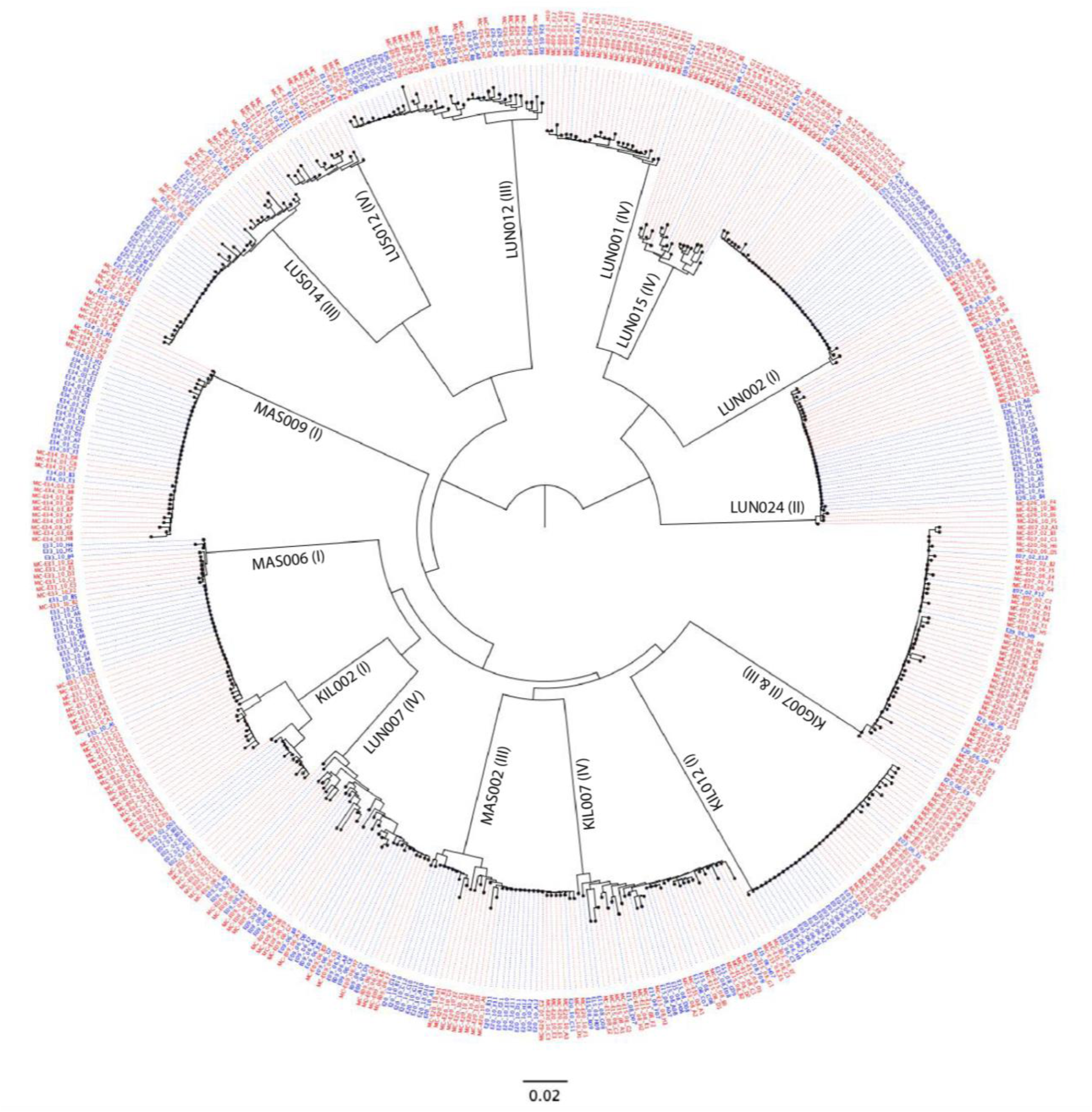
A phylogenetic tree showing relatedness of HIV-1 *env V1-V3* sequences generated from molecular cloning (red) and single genome amplification (blue). Labels “LUN001 (IV)”, “LUN015 (IV)”, “LUN002 (I)”, “LUN024 (III)”, and “LUN007 (IV)” indicate samples from the clinical cohort, and the remaining samples are from the research cohort. Values in parentheses indicate visit number.

**Table 2.**
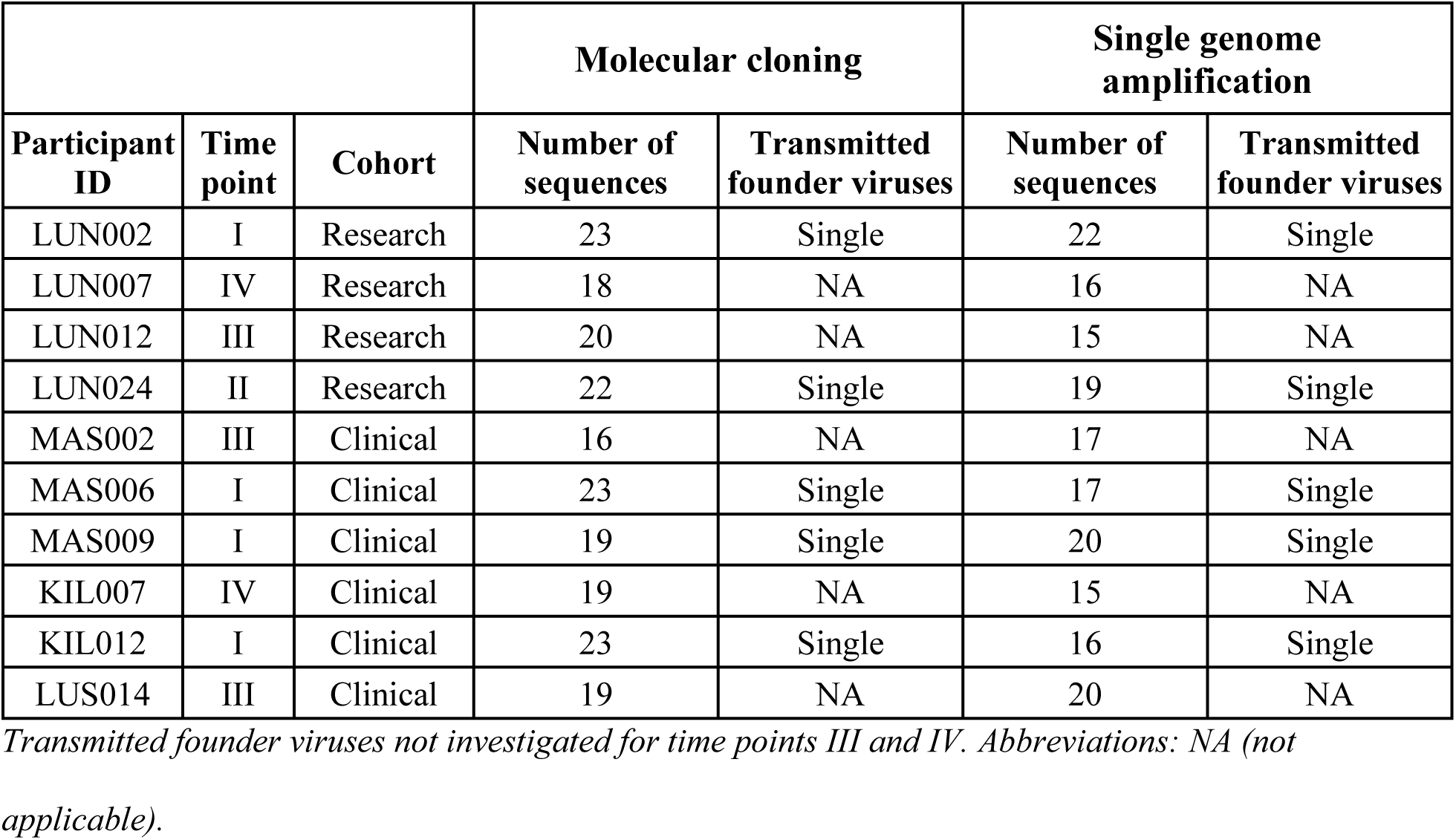
Samples involved in the comparison between single genome amplification and molecular cloning.

|  |  |  | **Molecular cloning** |  | **Single genome amplification** |  |
| --- | --- | --- | --- | --- | --- | --- |
| **Participant ID** | **Time point** | **Cohort** | **Number of sequences** | **Transmitted founder viruses** | **Number of sequences** | **Transmitted founder viruses** |
| LUN002 | I | Research | 23 | Single | 22 | Single |
| LUN007 | IV | Research | 18 | NA | 16 | NA |
| LUN012 | III | Research | 20 | NA | 15 | NA |
| LUN024 | II | Research | 22 | Single | 19 | Single |
| MAS002 | III | Clinical | 16 | NA | 17 | NA |
| MAS006 | I | Clinical | 23 | Single | 17 | Single |
| MAS009 | I | Clinical | 19 | Single | 20 | Single |
| KIL007 | IV | Clinical | 19 | NA | 15 | NA |
| KIL012 | I | Clinical | 23 | Single | 16 | Single |
| LUS014 | III | Clinical | 19 | NA | 20 | NA |

3 *Transmitted founder viruses not investigated for time points III and IV. Abbreviations: NA (not* 4 *applicable).*

The diversity analysis indicated consistently higher HIV-1 sequence diversity in the MC sequences compared with the corresponding SGA sequences (Figure 4). The overall mean diversities between MC and SGA sequence data were 0.009±0.007 and 0.006±0.006 nucleotide substitutions/site, respectively (p<0.001). Furthermore, higher diversity among MC relative to SGA sequences was observed among sequences from both research (0.010±0.009 vs. 0.007±0.008 nucleotide substitutions/site, p=0.022) and the clinical (0.008±0.006 vs. 0.005±0.005 nucleotide substitutions/site, p=0.021) cohorts. However, diversity estimates did not differ between research and clinical cohorts (0.008±0.008 vs 0.007±0.005 nucleotide substitutions/site, p=0.57).

**Figure 4.**
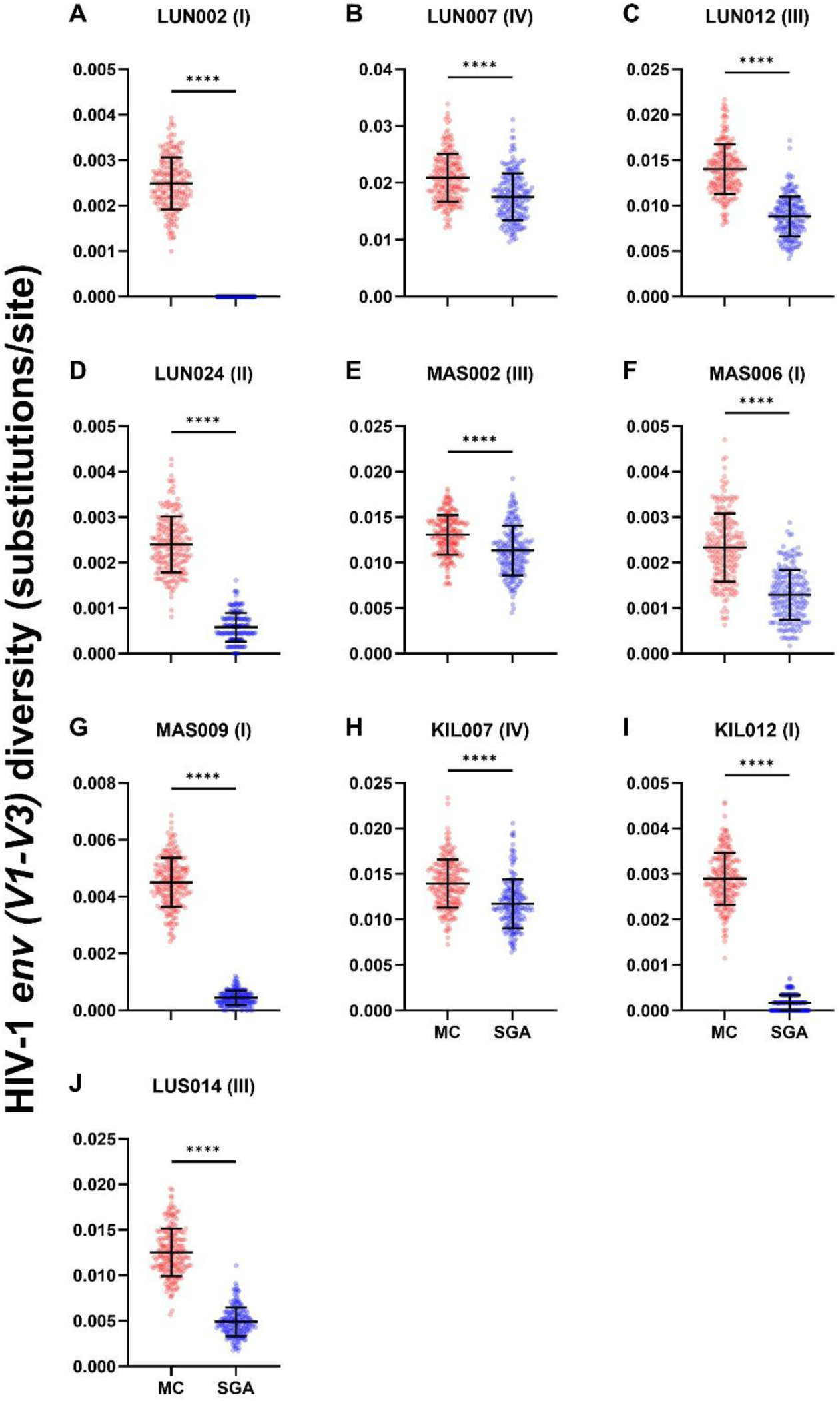
HIV-1 *env V1-V3* diversity compared between molecular cloning (red) and single genome amplification (blue) methods. Participants from the (A-F) clinical and (G-P) research cohorts are shown. Values in parentheses indicate visit number. Abbreviations: MC (molecular cloning), SGA (single genome amplification).

Within participants selected for SGA and MC comparisons, five samples were either time point I or II and thus eligible for TFV quantification (Table 2). All five samples were found to have a single TFV regardless of SGA or MC method used (Figure S4).

### Comparison of evolutionary rate between SGA and MC

Non-zero evolutionary rates were detected in all samples (Figure 5). Furthermore, there was significant between-patient variability in the HIV-1 evolutionary rates, regardless of whether the estimates were determined using SGA or MC methods. However, within-patient HIV-1 evolutionary rate estimates between SGA and MC methods were comparable (mean±SD 0.014±0.009 vs. 0.014±0.006 substitutions/site/year for SGA and MC data, respectively, p=0.673).

**Figure 5.**
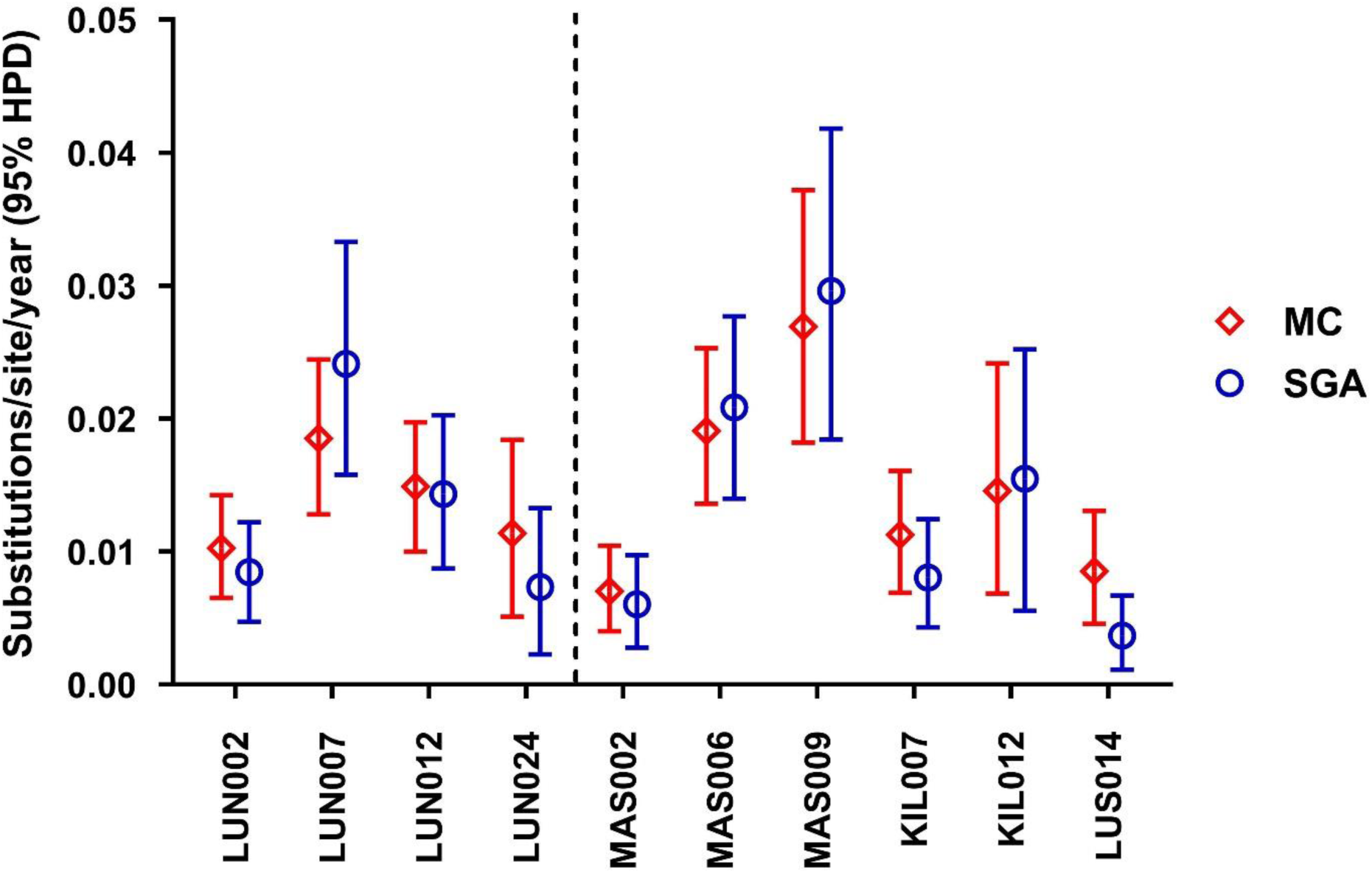
HIV-1 *env V1-V3* evolutionary rates compared between molecular cloning (red) and single genome amplification (blue) methods. Dotted line separates clinical (left) from research (right) cohort participants. Abbreviations: MC (molecular cloning), SGA (single genome amplification).

## DISCUSSION

Here, we present a comparison between SGA and MC methods to measure HIV-1 sequence diversity and evolutionary rate in participants with AHI. We corroborate previous findings by demonstrating that measured HIV-1 sequence diversity is higher by MC relative to SGA^6^. We also show for the first time that measurements of within-host evolutionary rates are comparable between the two methods, suggesting that errors intrinsic to the MC method do not propagate into evolutionary rate. These analyses were done using samples from two sources with differing sample quality, and these findings were true independent of sample quality and cohort type. Hence, for circumstances in which samples cannot be processed by SGA due to unavoidable variability in sample quality, use of MC can be justified to supplement measurements of evolutionary rate.

Several studies have directly compared SGA and MC methods. Salazar-Gonzalez *et al.* reported higher diversity and prevalence of recombinant sequences by MC compared to SGA of HIV-1 *env* sequences within two participants^6^. In a later study, Jordan *et al.* compared the two techniques in 17 participants to investigate inter-individual diversity in HIV-1 *pro-pol* sequences and found no differences in diversity^37^. However, the authors noted that a sufficient number of analysed sequences is necessary to detect low prevalence quasispecies^38^, and the necessary number of sequences to detect differences between the techniques is likely greater using *pro-pol* than *env*, given that the latter is more variable and less conserved. We used our dataset to empirically model the necessary number of sequences for stable diversity estimates and found eight sequences to be the approximate threshold. However, this is based on the within-host diversity of the population we sampled. It is possible that different thresholds exist for different populations, such as a chronic infection setting, in which diversity is likely to be higher.

Contending with poorer sample quality often accompanies studies using historical cohorts. In our analysis, research cohort samples returned more sequences, had fewer rejected sequences, and better concordance between qPCR measurement and pVL compared to clinical cohort samples. We observed that for several clinical cohort samples, historical HIV-1 pVL at visit could not be detected by qPCR when assayed years later. Although the clinical cohort tended to have older samples, we could not detect a relationship between sample age and quality in sensitivity analyses restricted to newer samples. It is possible that storage conditions (−80°C for research cohort and −20°C for clinical cohort samples) or cohort effects such as differences in sample handling/processing also played a role in sample degradation. Despite this, we show that the variability in diversity estimates between SGA and MC methods outweighs the variability between cohorts, suggesting that the heterogeneity in the samples used did not strongly affect the comparison.

Several studies have incorporated qPCR or digital PCR quantification of extracted HIV-1 genomes in the SGA workflow to inform the dilution to a single genome per sequencing reaction^24,39^. We expand on the utility of the qPCR data by describing the agreement between qPCR data and pVL at visit as a proxy of sample quality. We demonstrate that suboptimal sample quality in older samples is more likely to be reflected in divergence between HIV-1 qPCR and pVL concentrations. Thus, for studies in which variable sample quality is likely to lead to failed SGA sequencing, qPCR and pVL concordance could be used to screen for samples that would benefit from MC supplementation. Conversely, for studies in which sample quality is consistently high, the variability in pVL measurements is largely captured by qPCR. This suggests that the former may be used to inform SGA serial dilutions, bypassing the qPCR step. However, the performance and reliability of this warrants an empiric evaluation.

A variety of next generation sequencing platforms have become commonplace in HIV-1 genomics research^40,41^, including studies of acute infection^40^. However, these techniques often have limited read length and are vulnerable to sequencing errors and sample contamination compared to Sanger sequencing^42^. They also rely on *in silico* methods to stitch together sequencing fragments to form the original genomes. These limitations may constrain the usefulness of next generation techniques in diversity/evolution studies. Large fragment single molecule sequencing could, however, be a uniquely viable substitute for SGA, and has been used to study full-length *env* sequences^43^. Although it is applicable for HIV-1 diversity/evolution studies, it remains vulnerable to recombination error and robustness with low starting material has yet to be determined.

In our sequence analysis pipeline, we identify and exclude putative recombinant sequences from downstream analysis. Given the propensity of the MC method to introduce recombination errors^21,22^, it is critical to delineate *in vivo* from PCR-induced recombination. However, our data suggest that this does not explain the entirety of the error intrinsic to the MC method, as even among samples that have passed screening for putative recombinants, estimated diversity was higher than the SGA data. Indeed, after screening for recombinant sequences, two samples in our analysis had no sequence diversity when measured by SGA, but several nucleotide substitutions were detected when measured by MC.

Our analysis benefits from a range of sample quality characteristic of a large, multi-site AHI cohort study. We directly compare SGA and MC methods on the same samples to truly describe how MC measurements of evolutionary rate can supplement SGA data in this context. The low number of SGA reads in several samples was realistic, and we show that although increasing the number of sequences did not affect diversity estimates, sampling sequences fewer than a threshold of approximately eight sequences often underestimated diversity estimates. This threshold did not appear to be different between SGA and MC data, with the overestimation of diversity using MC seeming to be present across the range of sequences used, even down to two sequences. This is likely because the error is introduced in the PCR amplification stage and is already present prior to clone selection and sequencing. This suggests that error inherent to the MC method is constant and not dependent on the number of sequences generated.

The value of human cohort studies that span a long period of time or across different settings is indisputable. However, these expansive enterprises often rely on analyses of historic samples and/or samples collected from various sites, leading to unavoidable variability in sample quality. This work is an effort to strike the balance between maintaining the quality of data and conserving the value of precious samples.

## Data Availability

All data produced in the present study are available upon reasonable request to the authors.

## Acknowledgements

We are grateful to IAVI for supporting HIV-1 research studies and capacity building initiatives in Kenya, Uganda, Rwanda and Zambia. We are also grateful to staff and participants from IAVI sites in Africa and from the Department of Infectious Diseases at Skåne University Hospital in Sweden, without whom this work would not have been possible. We acknowledge the following people for their generous contributions and support of this project: Jan Albert (Karolinska Institute, Sweden), and Bengt Löfgren, Bertil Christensson, and Karin Behrens (all from SUS Skåne, Sweden). The report is published with permission from the Kenya Medical Research Institute (KEMRI).

## Author contributions

J.E and A.S.H conceptualised the study and designed the analysis plan. A.Y.Y.H, A.S.H, and L.L carried out the analyses, and A.Y.Y.H and A.S.H prepared the drafted manuscript. W.K, E.K, M.P, A.K, P.K, J.T, S.A, E.H, and J.G generated the data on the cohorts. W.K, E.K, M.P, A.K, P.K, S.A, E.H, J.G, S.R-J, E.J.S, A.S.H and J.E reviewed the manuscript and provided feedback. All authors approved the final draft of the manuscript for submission.

## REFERENCES

1. Zhu T, Mo H, Wang N, Nam DS, Cao Y, Koup RA, Ho DD. Genotypic and phenotypic characterization of HIV-1 patients with primary infection. Science. 1993;261(5125):1179–1181. doi:10.1126/science.8356453

2. Delwart E, Magierowska M, Royz M, Foley B, Peddada L, Smith R, Heldebrant C, Conrad A, Busch M. Homogeneous quasispecies in 16 out of 17 individuals during very early HIV-1 primary infection. AIDS. 2002;16(2).

3. Learn GH, Muthui D, Brodie SJ, Zhu T, Diem K, Mullins JI, Corey L. Virus population homogenization following acute human immunodeficiency virus type 1 infection. J Virol. 2002;76(23):11953–11959. doi:10.1128/jvi.76.23.11953-11959.2002

4. Keele BF, Giorgi EE, Salazar-Gonzalez JF, Decker JM, Pham KT, Salazar MG, Sun C, Grayson T, Wang S, Li H, Wei X, Jiang C, Kirchherr JL, Gao F, Anderson JA, Ping L-H, Swanstrom R, Tomaras GD, Blattner WA, Goepfert PA, Kilby JM, Saag MS, Delwart EL, Busch MP, Cohen MS, Montefiori DC, Haynes BF, Gaschen B, Athreya GS, Lee HY, Wood N, Seoighe C, Perelson AS, Bhattacharya T, Korber BT, Hahn BH, Shaw GM. Identification and characterization of transmitted and early founder virus envelopes in primary HIV-1 infection. Proc Natl Acad Sci U S A. 2008;105(21):7552–7557. doi:10.1073/pnas.0802203105

5. Derdeyn CA, Decker JM, Bibollet-Ruche F, Mokili JL, Muldoon M, Denham SA, Heil ML, Kasolo F, Musonda R, Hahn BH, Shaw GM, Korber BT, Allen S, Hunter E. Envelope-Constrained Neutralization-Sensitive HIV-1 After Heterosexual Transmission. Science. 2004;303(5666):2019–2022. doi:10.1126/science.1093137

6. Salazar-Gonzalez JF, Bailes E, Pham KT, Salazar MG, Guffey MB, Keele BF, Derdeyn CA, Farmer P, Hunter E, Allen S, Manigart O, Mulenga J, Anderson JA, Swanstrom R, Haynes BF, Athreya GS, Korber BTM, Sharp PM, Shaw GM, Hahn BH. Deciphering human immunodeficiency virus type 1 transmission and early envelope diversification by single-genome amplification and sequencing. J Virol. 2008;82(8):3952–3970. doi:10.1128/JVI.02660-07

7. Salazar-Gonzalez JF, Salazar MG, Keele BF, Learn GH, Giorgi EE, Li H, Decker JM, Wang S, Baalwa J, Kraus MH, Parrish NF, Shaw KS, Guffey MB, Bar KJ, Davis KL, Ochsenbauer-Jambor C, Kappes JC, Saag MS, Cohen MS, Mulenga J, Derdeyn CA, Allen S, Hunter E, Markowitz M, Hraber P, Perelson AS, Bhattacharya T, Haynes BF, Korber BT, Hahn BH, Shaw GM. Genetic identity, biological phenotype, and evolutionary pathways of transmitted/founder viruses in acute and early HIV-1 infection. J Exp Med. 2009;206(6):1273–1289. doi:10.1084/jem.20090378

8. Abrahams M-R, Anderson JA, Giorgi EE, Seoighe C, Mlisana K, Ping L-H, Athreya GS, Treurnicht FK, Keele BF, Wood N, Salazar-Gonzalez JF, Bhattacharya T, Chu H, Hoffman I, Galvin S, Mapanje C, Kazembe P, Thebus R, Fiscus S, Hide W, Cohen MS, Karim SA, Haynes BF, Shaw GM, Hahn BH, Korber BT, Swanstrom R, Williamson C. Quantitating the multiplicity of infection with human immunodeficiency virus type 1 subtype C reveals a non-poisson distribution of transmitted variants. J Virol. 2009;83(8):3556–3567. doi:10.1128/JVI.02132-08

9. Joseph SB, Swanstrom R, Kashuba ADM, Cohen MS. Bottlenecks in HIV-1 transmission: insights from the study of founder viruses. Nat Rev Microbiol. 2015;13(7):414–425. doi:10.1038/nrmicro3471

10. Carlson JM, Schaefer M, Monaco DC, Batorsky R, Claiborne DT, Prince J, Deymier MJ, Ende ZS, Klatt NR, DeZiel CE, Lin T-H, Peng J, Seese AM, Shapiro R, Frater J, Ndung’u T, Tang J, Goepfert P, Gilmour J, Price MA, Kilembe W, Heckerman D, Goulder PJR, Allen TM, Allen S, Hunter E. HIV transmission. Selection bias at the heterosexual HIV-1 transmission bottleneck. Science. 2014;345(6193):1254031. doi:10.1126/science.1254031

11. Shankarappa R, Margolick JB, Gange SJ, Rodrigo AG, Upchurch D, Farzadegan H, Gupta P, Rinaldo CR, Learn GH, He X, Huang XL, Mullins JI. Consistent viral evolutionary changes associated with the progression of human immunodeficiency virus type 1 infection. J Virol. 1999;73(12):10489–10502. doi:10.1128/JVI.73.12.10489-10502.1999

12. Lemey P, Kosakovsky Pond SL, Drummond AJ, Pybus OG, Shapiro B, Barroso H, Taveira N, Rambaut A. Synonymous substitution rates predict HIV disease progression as a result of underlying replication dynamics. PLoS Comput Biol. 2007;3(2):e29. doi:10.1371/journal.pcbi.0030029

13. Esbjörnsson J, Månsson F, Kvist A, Isberg P-E, Nowroozalizadeh S, Biague AJ, da Silva ZJ, Jansson M, Fenyö EM, Norrgren H, Medstrand P. Inhibition of HIV-1 disease progression by contemporaneous HIV-2 infection. N Engl J Med. 2012;367(3):224–232. doi:10.1056/NEJMoa1113244

14. Maldarelli F, Kearney M, Palmer S, Stephens R, Mican J, Polis MA, Davey RT, Kovacs J, Shao W, Rock-Kress D, Metcalf JA, Rehm C, Greer SE, Lucey DL, Danley K, Alter H, Mellors JW, Coffin JM. HIV populations are large and accumulate high genetic diversity in a nonlinear fashion. J Virol. 2013;87(18):10313–10323. doi:10.1128/JVI.01225-12

15. Simmonds P, Balfe P, Ludlam CA, Bishop JO, Brown AJ. Analysis of sequence diversity in hypervariable regions of the external glycoprotein of human immunodeficiency virus type 1. J Virol. 1990;64(12):5840–5850. doi:10.1128/JVI.64.12.5840-5850.1990

16. Goonetilleke N, Liu MKP, Salazar-Gonzalez JF, Ferrari G, Giorgi E, Ganusov V V, Keele BF, Learn GH, Turnbull EL, Salazar MG, Weinhold KJ, Moore S, Letvin N, Haynes BF, Cohen MS, Hraber P, Bhattacharya T, Borrow P, Perelson AS, Hahn BH, Shaw GM, Korber BT, McMichael AJ. The first T cell response to transmitted/founder virus contributes to the control of acute viremia in HIV-1 infection. J Exp Med. 2009;206(6):1253–1272. doi:10.1084/jem.20090365

17. Ho Y-C, Shan L, Hosmane NN, Wang J, Laskey SB, Rosenbloom DIS, Lai J, Blankson JN, Siliciano JD, Siliciano RF. Replication-competent noninduced proviruses in the latent reservoir increase barrier to HIV-1 cure. Cell. 2013;155(3):540–551. doi:10.1016/j.cell.2013.09.020

18. Jones BR, Miller RL, Kinloch NN, Tsai O, Rigsby H, Sudderuddin H, Shahid A, Ganase B, Brumme CJ, Harris M, Poon AFY, Brockman MA, Fromentin R, Chomont N, Joy JB, Brumme ZL. Genetic Diversity, Compartmentalization, and Age of HIV Proviruses Persisting in CD4(+) T Cell Subsets during Long-Term Combination Antiretroviral Therapy. J Virol. 2020;94(5). doi:10.1128/JVI.01786-19

19. Poss M, Martin HL, Kreiss JK, Granville L, Chohan B, Nyange P, Mandaliya K, Overbaugh J. Diversity in virus populations from genital secretions and peripheral blood from women recently infected with human immunodeficiency virus type 1. J Virol. 1995;69(12):8118–8122. doi:10.1128/JVI.69.12.8118-8122.1995

20. McCutchan FE, Hoelscher M, Tovanabutra S, Piyasirisilp S, Sanders-Buell E, Ramos G, Jagodzinski L, Polonis V, Maboko L, Mmbando D, Hoffmann O, Riedner G, von Sonnenburg F, Robb M, Birx DL. In-depth analysis of a heterosexually acquired human immunodeficiency virus type 1 superinfection: evolution, temporal fluctuation, and intercompartment dynamics from the seronegative window period through 30 months postinfection. J Virol. 2005;79(18):11693–11704. doi:10.1128/JVI.79.18.11693-11704.2005

21. Yang YL, Wang G, Dorman K, Kaplan AH. Long polymerase chain reaction amplification of heterogeneous HIV type 1 templates produces recombination at a relatively high frequency. AIDS Res Hum Retroviruses. 1996;12(4):303–306. doi:10.1089/aid.1996.12.303

22. Fang G, Zhu G, Burger H, Keithly JS, Weiser B. Minimizing DNA recombination during long RT-PCR. J Virol Methods. 1998;76(1-2):139–148. doi:10.1016/s0166-0934(98)00133-5

23. Liu S-L, Rodrigo AG, Shankarappa R, Learn GH, Hsu L, Davidov O, Zhao LP, Mullins JI. HIV Quasispecies and Resampling. Science. 1996;273(5274):415–416. doi:10.1126/science.273.5274.415

24. Palmer S, Kearney M, Maldarelli F, Halvas EK, Bixby CJ, Bazmi H, Rock D, Falloon J, Davey RTJ, Dewar RL, Metcalf JA, Hammer S, Mellors JW, Coffin JM. Multiple, linked human immunodeficiency virus type 1 drug resistance mutations in treatment-experienced patients are missed by standard genotype analysis. J Clin Microbiol. 2005;43(1):406–413. doi:10.1128/JCM.43.1.406-413.2005

25. Hassan AS, Hare J, Kamini G, Yindom LM, Kamali A, Karita E, Kilemba W, Price MA, Borrow P, Bjorkman P, Albert J, Kaleebu P, Allan S, Fast P, Hunter E, Gilmour J, Ndung’u T, Rowland-Jones S, Sanders EJ, Esbjornsson J. A35 Viral evolution and innate immune responses during acute HIV-1 infection and their association with disease pathogenesis. Virus Evol. 2017;3(Suppl 1). doi:10.1093/ve/vew036.034

26. Hassan AS, Hare J, Gounder K, Nazziwa J, Karlson S, Olsson L, Streatfield C, Kamali A, Karita E, Kilembe W, Price MA, Borrow P, Björkman P, Kaleebu P, Allen S, Hunter E, Ndung’u T, Gilmour J, Rowland-Jones S, Esbjörnsson J, Sanders EJ. A Stronger Innate Immune Response During Hyperacute Human Immunodeficiency Virus Type 1 (HIV-1) Infection Is Associated With Acute Retroviral Syndrome. Clin Infect Dis. 2021;73(5):832–841. doi:10.1093/cid/ciab139

27. Nazziwa J, Freyhult E, Hong M-G, Johansson E, Årman F, Hare J, Gounder K, Rezeli M, Mohanty T, Kjellström S, Kamali A, Karita E, Kilembe W, Price MA, Kaleebu P, Allen S, Hunter E, Ndung’u T, Gilmour J, Rowland-Jones SL, Sanders E, Hassan AS, Esbjörnsson J. Dynamics of the blood plasma proteome during hyperacute HIV-1 infection. Nat Commun. 2024;15(1):10593. doi:10.1038/s41467-024-54848-0

28. Kamali A, Price MA, Lakhi S, Karita E, Inambao M, Sanders EJ, Anzala O, Latka MH, Bekker L-G, Kaleebu P, Asiki G, Ssetaala A, Ruzagira E, Allen S, Farmer P, Hunter E, Mutua G, Makkan H, Tichacek A, Brill IK, Fast P, Stevens G, Chetty P, Amornkul PN, Gilmour J. Creating an African HIV clinical research and prevention trials network: HIV prevalence, incidence and transmission. PLoS One. 2015;10(1):e0116100. doi:10.1371/journal.pone.0116100

29. Fiebig EW, Wright DJ, Rawal BD, Garrett PE, Schumacher RT, Peddada L, Heldebrant C, Smith R, Conrad A, Kleinman SH, Busch MP. Dynamics of HIV viremia and antibody seroconversion in plasma donors: implications for diagnosis and staging of primary HIV infection. AIDS. 2003;17(13):1871–1879. doi:10.1097/00002030-200309050-00005

30. Price MA, Kilembe W, Ruzagira E, Karita E, Inambao M, Sanders EJ, Anzala O, Allen S, Edward VA, Kaleebu P, Fast PE, Rida W, Kamali A, Hunter E, Tang J, Lakhi S, Mutua G, Bekker LG, Abu-Baker G, Tichacek A, Chetty P, Latka MH, Maenetje P, Makkan H, Hare J, Kibengo F, Priddy F, Landais E, Chinyenze K, Gilmour J. Cohort Profile: IAVI’s HIV epidemiology and early infection cohort studies in Africa to support vaccine discovery. Int J Epidemiol. 2021;50(1):29–30. doi:10.1093/ije/dyaa100

31. Esbjörnsson J, Mild M, Månsson F, Norrgren H, Medstrand P. HIV-1 molecular epidemiology in Guinea-Bissau, West Africa: origin, demography and migrations. PLoS One. 2011;6(2):e17025. doi:10.1371/journal.pone.0017025

32. Esbjörnsson J, Månsson F, Martínez-Arias W, Vincic E, Biague AJ, da Silva ZJ, Fenyö EM, Norrgren H, Medstrand P. Frequent CXCR4 tropism of HIV-1 subtype A and CRF02_AG during late-stage disease--indication of an evolving epidemic in West Africa. Retrovirology. 2010;7:23. doi:10.1186/1742-4690-7-23

33. Mild M, Esbjörnsson J, Fenyö EM, Medstrand P. Frequent intrapatient recombination between human immunodeficiency virus type 1 R5 and X4 envelopes: implications for coreceptor switch. J Virol. 2007;81(7):3369–3376. doi:10.1128/JVI.01295-06

34. Leitner T, Korovina G, Marquina S, Smolskaya T, Albert J. Molecular epidemiology and MT-2 cell tropism of Russian HIV type 1 variant. AIDS Res Hum Retroviruses. 1996;12(17):1595–1603. doi:10.1089/aid.1996.12.1595

35. Bruen TC, Philippe H, Bryant D. A simple and robust statistical test for detecting the presence of recombination. Genetics. 2006;172(4):2665–2681. doi:10.1534/genetics.105.048975

36. Palm AA, Lemey P, Jansson M, Månsson F, Kvist A, Szojka Z, Biague A, da Silva ZJ, Rowland-Jones SL, Norrgren H, Esbjörnsson J, Medstrand P. Low Postseroconversion CD4(+) T-cell Level Is Associated with Faster Disease Progression and Higher Viral Evolutionary Rate in HIV-2 Infection. MBio. 2019;10(1). doi:10.1128/mBio.01245-18

37. Jordan MR, Kearney M, Palmer S, Shao W, Maldarelli F, Coakley EP, Chappey C, Wanke C, Coffin JM. Comparison of standard PCR/cloning to single genome sequencing for analysis of HIV-1 populations. J Virol Methods. 2010;168(1-2):114–120. doi:10.1016/j.jviromet.2010.04.030

38. Eric L, Yifan L, Hongjun B, Phuc P, Morgane R. HIV-1 Gag, Pol, and Env diversified with limited adaptation since the 1980s. MBio. 2024;15(3):e01749–23. doi:10.1128/mbio.01749-23

39. Lee GQ, Reddy K, Einkauf KB, Gounder K, Chevalier JM, Dong KL, Walker BD, Yu XG, Ndung’u T, Lichterfeld M. HIV-1 DNA sequence diversity and evolution during acute subtype C infection. Nat Commun. 2019;10(1):2737. doi:10.1038/s41467-019-10659-2

40. Liang B, Luo M, Scott-Herridge J, Semeniuk C, Mendoza M, Capina R, Sheardown B, Ji H, Kimani J, Ball BT, Van Domselaar G, Graham M, Tyler S, Jones SJM, Plummer FA. A comparison of parallel pyrosequencing and sanger clone-based sequencing and its impact on the characterization of the genetic diversity of HIV-1. PLoS One. 2011;6(10):e26745. doi:10.1371/journal.pone.0026745

41. Kijak GH, Sanders-Buell E, Pham P, Harbolick EA, Oropeza C, O’Sullivan AM, Bose M, Beckett CG, Milazzo M, Robb ML, Peel SA, Scott PT, Michael NL, Armstrong AW, Kim JH, Brett-Major DM, Tovanabutra S. Next-generation sequencing of HIV-1 single genome amplicons. Biomol Detect Quantif. 2019;17:100080. doi:10.1016/j.bdq.2019.01.002

42. Brumme CJ, Poon AFY. Promises and pitfalls of Illumina sequencing for HIV resistance genotyping. Virus Res. 2017;239:97–105. doi:10.1016/j.virusres.2016.12.008

43. Laird Smith M, Murrell B, Eren K, Ignacio C, Landais E, Weaver S, Phung P, Ludka C, Hepler L, Caballero G, Pollner T, Guo Y, Richman D, Poignard P, Paxinos EE, Kosakovsky Pond SL, Smith DM. Rapid Sequencing of Complete env Genes from Primary HIV-1 Samples. Virus Evol. 2016;2(2):vew018. doi:10.1093/ve/vew018

