## Supplementary material for "Single genome amplification and molecular cloning of HIV-1 populations in acute HIV-1 infection: implications for studies on HIV-1 diversity and evolutionary rate"

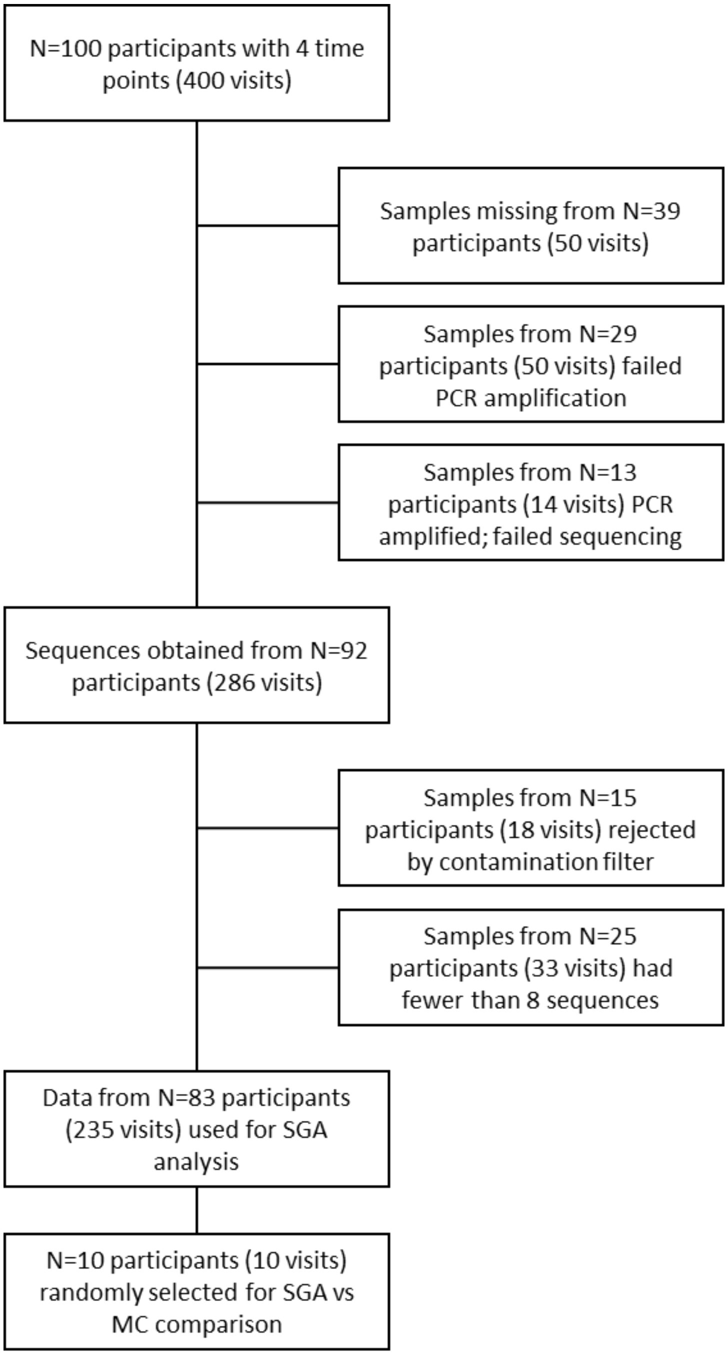


**Figure S1.** Inclusion criteria of participants and samples for single genome amplification sequencing and downstream analysis. Abbreviations: SGA (single genome amplification).

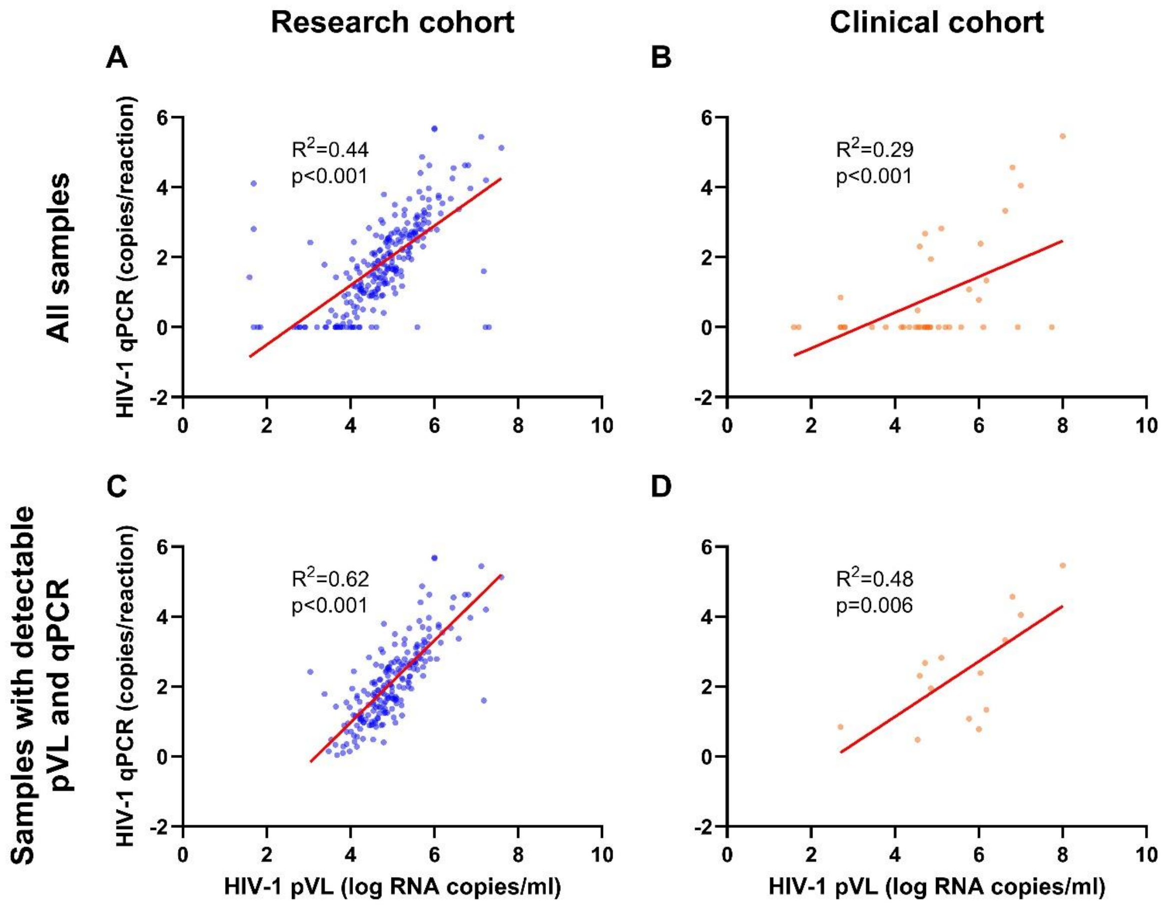


**Figure S2.** Associations between measurements of HIV-1 RNA at visit (HIV-1 pVL) and during sequencing (HIV-1 qPCR). All samples in the (A) research and (B) clinical cohorts are shown. A subset of samples with detectable RNA both at visit and during sequencing in (C) research and (D) clinical cohorts are shown. The red line indicates line of best fit, and Pearson's $R^2$ and p-values are shown. Abbreviations: pVL (plasma viral load), qPCR (quantitative polymerase chain reaction).

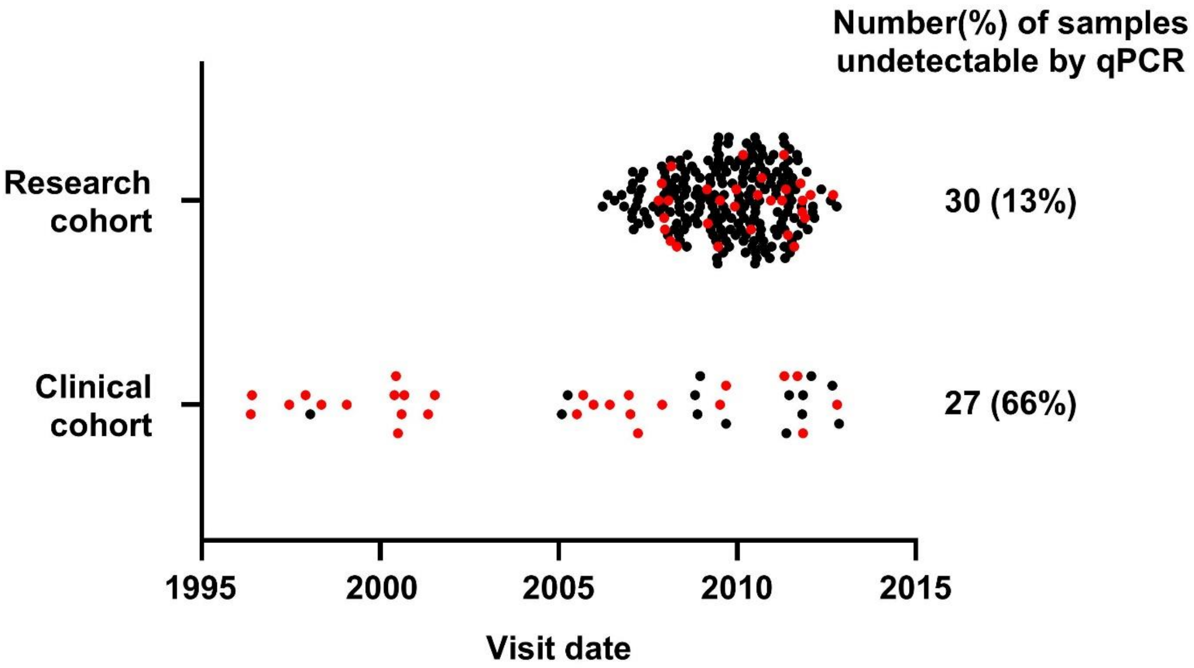


**Figure S3.** Visit dates of participants in the research and clinical cohorts. Red dots indicate detectable HIV-1 pVL at visit but undetectable HIV RNA by qPCR during sequencing. Abbreviations: pVL (plasma viral load), qPCR (quantitative polymerase chain reaction).

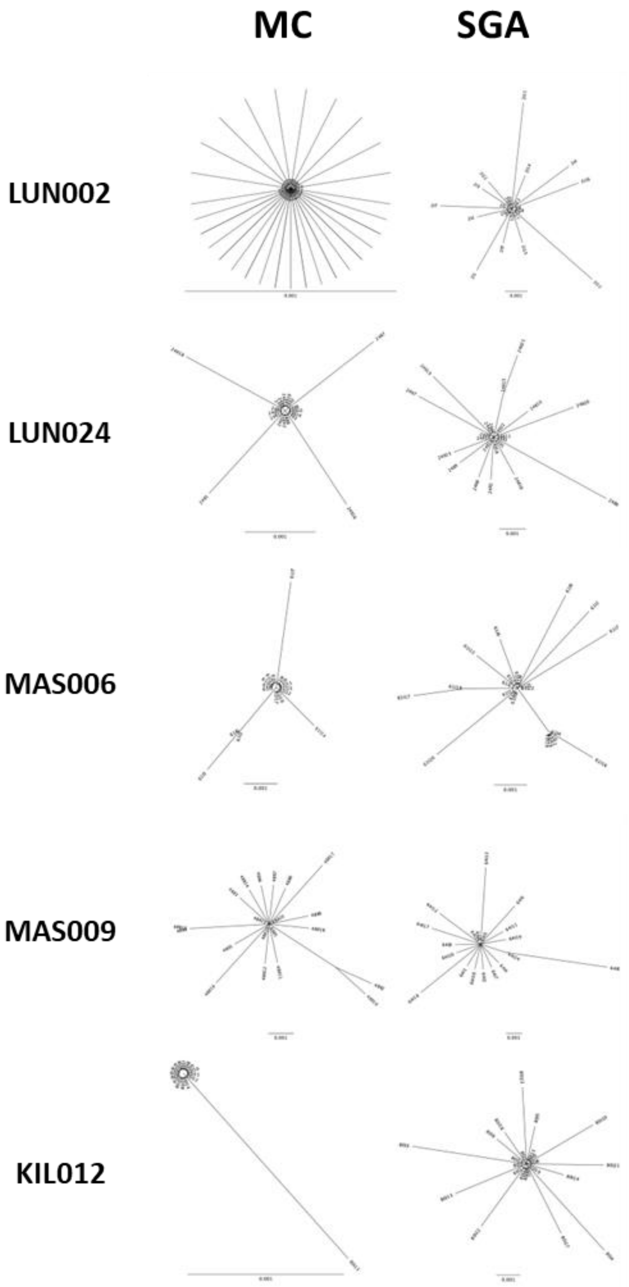


**Figure S4.** Neighbour-joining phylogenetic trees characterizing monophyletic lineage in each of the five participants eligible for transmitted founder virus quantification using either MC or SGA sequences. Abbreviations: MC (molecular cloning), SGA (single genome amplification).
